# Call Me Dr. Ishmael: Trends in Electronic Health Record Notes Available at ED Visits and Admissions

**DOI:** 10.1101/2024.02.23.24303213

**Authors:** Brian W. Patterson, Daniel J Hekman, Frank Liao, Azita Hamedani, Manish N. Shah, Majid Afshar

## Abstract

**Objective:** Numerous studies have identified information overload as a key issue for electronic health records (EHRs). This study describes the amount of text data across all notes available to emergency physicians in the EHR, trended over the time since EHR establishment.

**Materials and Methods:** We conducted a retrospective analysis of EHR data from a large healthcare system, examining the number of notes and corresponding number of total words and total tokens across all notes available to physicians during patient encounters in the emergency department (ED). We assessed the change in these metrics over a 17-year period between 2006 and 2023.

**Results:** The study cohort included 730,968 ED visits made by 293,559 unique patients and a total note count of 132,574,964. The median note count for all encounters in 2006 was 7 (IQR: 3 - 18), accounting for 1,894 words (IQR: 538 - 5,864). By the last full year of the study period in 2022, the median number of notes had grown to 380 (IQR: 93 - 1,008), representing 61,591 words (IQR: 13,621 - 174,152). Note and word counts were higher for admitted patients.

**Conclusion:** The volume of notes available for review by providers has increased by over 30-fold in the 17 years since the implementation of the EHR at a large health system. The task of reviewing these notes has become correspondingly more difficult. These data point to the critical need for new strategies and tools for filtering, synthesizing, and summarizing information to achieve the promise of the medical record.

## Background and Significance

Since the early 2000s, electronic health record systems (EHRs) have been nearly universally adopted in US hospital systems^1,2^. Widespread adoption of EHRs offers an unprecedented opportunity to improve care by collecting and storing clinical data in a format instantly accessible to clinicians.^3,4^ The EHR stores information from past visits, including notes, and makes this information instantly available to treating providers to inform care.^3–5^ This information is of critical importance to providers in the emergency department (ED) as they are often meeting patients for the first time and need to provide care for acute complications of complex, chronic medical conditions. Access to prior records allows providers to understand patients’ long term and recent health history, which can be difficult to obtain from patients themselves due to health literacy, chronic conditions such as dementia, or acute conditions such as delirium that prevent effective communication.

Often a provider’s first step prior to seeing a patient in the ED is performing a “chart biopsy”, in which they briefly look through a patient’s prior notes and other data to determine pertinent medical history which could impact the present visit.^6^ Given clinical demands on providers in the ED setting, this task is afforded a few minutes at most. However, as EHRs have grown in the scope of patient encounters documented, as well as time since deployment, the ability of any given provider to process these data effectively has become a concern in time-pressured settings such as the ED.^7–9^ In their brief patient encounters, providers in the ED are confronted with huge volumes of poorly organized information, which is difficult to synthesize and act upon at the bedside.^10,11^ The expectation for a provider to review available clinical data for a given patient has remained despite an increase in the amount of data available. “Missing” critical details of a patient’s medical history - including prior diagnoses, current medications, and/or recent lab values and imaging studies - is a significant concern. These concerns exist not only for emergency providers, but also for other providers tasked with acutely caring for patients they have no previous relationship with, including hospitalists and intensivists.

While prior work has suggested that the length of individual notes has increased,^12,13^ the overall quantity of text data available to clinicians in aggregate at a given clinical visit has not been evaluated.

## Objective

We sought to quantify the number and length of notes available to providers caring for patients in the ED, as measured by number of notes, total number of words among all notes, and total number of text tokens for use in a large language model (LLM). Additionally, we evaluated the trend in amount of data presented to providers over time since the inception of the EHR system. We hypothesized that the total number of notes as well as overall text data presented to clinicians has substantially increased. As subgroup analyses, we additionally examined patients who were admitted from the ED to general care beds and intensive care units (ICUs), with the hypothesis that these patients would be more complex and have more notes.

## Methods

### Patient Setting and Data Environment

We conducted a retrospective analysis of EHR data from quaternary care academic medical systems EHR. This EHR is used by an academic medical center that is also certified as a Level 1 Trauma. The same instance of the EHR is shared at an affiliate ED staffed by the same providers which opened in 2015, which was also included in this study. Combined, these two EDs now care for approximately 90,000 visits yearly. This study was IRB reviewed and granted an exemption as secondary research on existing data. The study cohort included patients aged 18 years and older at the time of an ED visit who presented to the ED between 3/10/2006 (the first year in which the EHR was deployed) and 1/31/2023. Note data were extracted from an EHR relational database (Epic Systems, Verona, WI). We included all notes available to providers (defined as physicians or advanced practice providers) during an ED encounter. This includes all information entered into the EHR as a note prior to an ED encounter, including provider-generated notes (progress notes, history and physicals, discharge summaries and consult notes), telephone notes, and nursing and other care team notes. These notes did not include text from lab, radiology, or other procedure reports which are filed as procedures or results and not included in the “notes” section of the EHR. All notes available up to the timestamp of the index ED encounter were included, but notes generated during the encounter were not. Only notes generated within the system were included (notes from other healthcare organizations may have been available for clinician review depending on the year, but these were not analyzed for this study). Notes and encounters deemed sensitive by our institution policy (e.g. those for patients who explicitly opted out of having their charts available for research) were excluded.

### Analysis

The unit of analysis was the patient chart as presented to the provider at the time of arrival to the ED. Data were collected at the encounter level such that one patient who had multiple ED encounters over the study period generated multiple datapoints, with the notes available at each encounter analyzed separately at each encounter. For instance, a patient may have had an encounter in 2015 where 10 notes were available, but another encounter in 2020 by which point 20 notes were available including the 10 available in 2015. This patient would result in two encounter data points included in the study, one with 10 and one with 20 notes available. This same patient may have been seen in 2010 while still a minor, but that encounter would be excluded from this study as an analysis point, however the notes generated during the 2010 encounter would be included as data for the 2015 and 2020 encounters.

Within each chart for each encounter, notes were individually parsed and then aggregated statistics were created by adding word and token counts from all available notes filed in the EHR prior to each ED arrival. Word count was generated by splitting text on spaces in Python (Python Software Foundation, 2019). We also generated token counts using a subword-based tokenizer (tiktoken^14^), known as byte pair encoding (BPE). This tokenizer approach is used for large language models (LLMs) like GPT for tokenization and is the approach for determining the token limits for LLMs.

Three metrics were calculated and trended by year over the study period: total number of notes, total number of words as unigrams, and total number of tokens. Median and interquartile ranges were created to quantify and compare the distribution over time. Results were graphically plotted using box plots, on a log scale to allow improved visualization of distributions. As a reference, we provided word counts of well-known English language works to benchmark patient note word counts. We additionally reported the patient demographics and characteristics as well as their associated note characteristics. All notes were processed in Python and the data were analyzed in R (R Development Core Team).

Demographic and other clinically relevant metrics for the patients presenting were abstracted and presented for informational purposes, however this study was not intended to examine the relationship between patient characteristics and chart size.

## Results

The study cohort included 730,968 ED visits made by 293,559 unique patients and a total note count of 132,574,964 over the 17-year study period. Figure 1 describes trends by year in the amount of note information available to providers over time by word count. Table 1 provides note counts, word length, and token count for select years, with Appendix A providing data for all years. The median note count for all encounters in 2006 was 7 (IQR: 3 - 18), accounting for 1,894 words (IQR: 538 - 5,864) and 3,236 tokens (IQR: 960 - 9,724). By the last full year of the study period, 2022, the median number of notes had grown to 380 (IQR: 93 - 1,008) notes, representing 61,591 (IQR: 13,621 - 174,152) words and 103,350 (IQR: 22,375 - 298,983)

**Table 1:** Notes, Words and Tokens Available at ED Encounters.

| Year | ED Encounters | Median (IQR) Previous Notes | Median (IQR) Previous Words | Median (IQR) Previous Tokens |
| --- | --- | --- | --- | --- |
| **All ED Encounters** |  |  |  |  |
| 2010 | 29,220 | 106 (25 - 308) | 13,179 (2,328 - 45,258) | 22,470 (3,907 - 78,514) |
| 2014 | 33,066 | 190 (45 - 543) | 27,322 (6,340 - 84,940) | 45,742 (10,432 - 146,021) |
| 2018 | 59,719 | 311 (75 - 841) | 51,147 (11,448 - 146,218) | 86,071 (18,834 - 251,322) |
| 2022 | 71,344 | 380 (93 - 1,008) | 61,591 (13,621 - 174,152) | 103,350 (22,375 - 298,983) |
| **ED Encounters Resulting in General Care Admission** |  |  |  |  |
| 2010 | 5,568 | 268 (82 - 626) | 40,920 (10,291 - 106,590) | 71,246 (17,752 - 190,243) |
| 2014 | 4,643 | 412 (116 - 1,025) | 67,970 (17,260 - 176,345) | 117,754 (29,468 - 312,853) |
| 2018 | 13,103 | 564 (162 - 1,356) | 100,763 (27,182 - 248,684) | 172,471 (45,854 - 435,658) |
| 2022 | 14,383 | 675 (202 - 1,636) | 117,906 (31,010 - 301,868) | 203,741 (52,154 - 529,416) |
| **ED Encounters Resulting in IMC/ICU Admissions** |  |  |  |  |
| 2010 | 726 | 243 (60 - 659) | 36,260 (5,833 - 118,834) | 63,677 (10,076 - 212,672) |
| 2014 | 754 | 372 (72 - 988) | 61,493 (7,592 - 183,691) | 106,934 (12,580 - 328,589) |
| 2018 | 1,879 | 473 (78 - 1,370) | 84,802 (10,164 - 262,356) | 145,940 (17,294 - 470,318) |
| 2022 | 2,489 | 560 (100 - 1,652) | 95,998 (12,364 - 316,354) | 164,848 (20,869 - 554,417) |

Note:
IMC: Intermediate Care Unit; ICU: Intensive Care Unit

**Figure 1:**
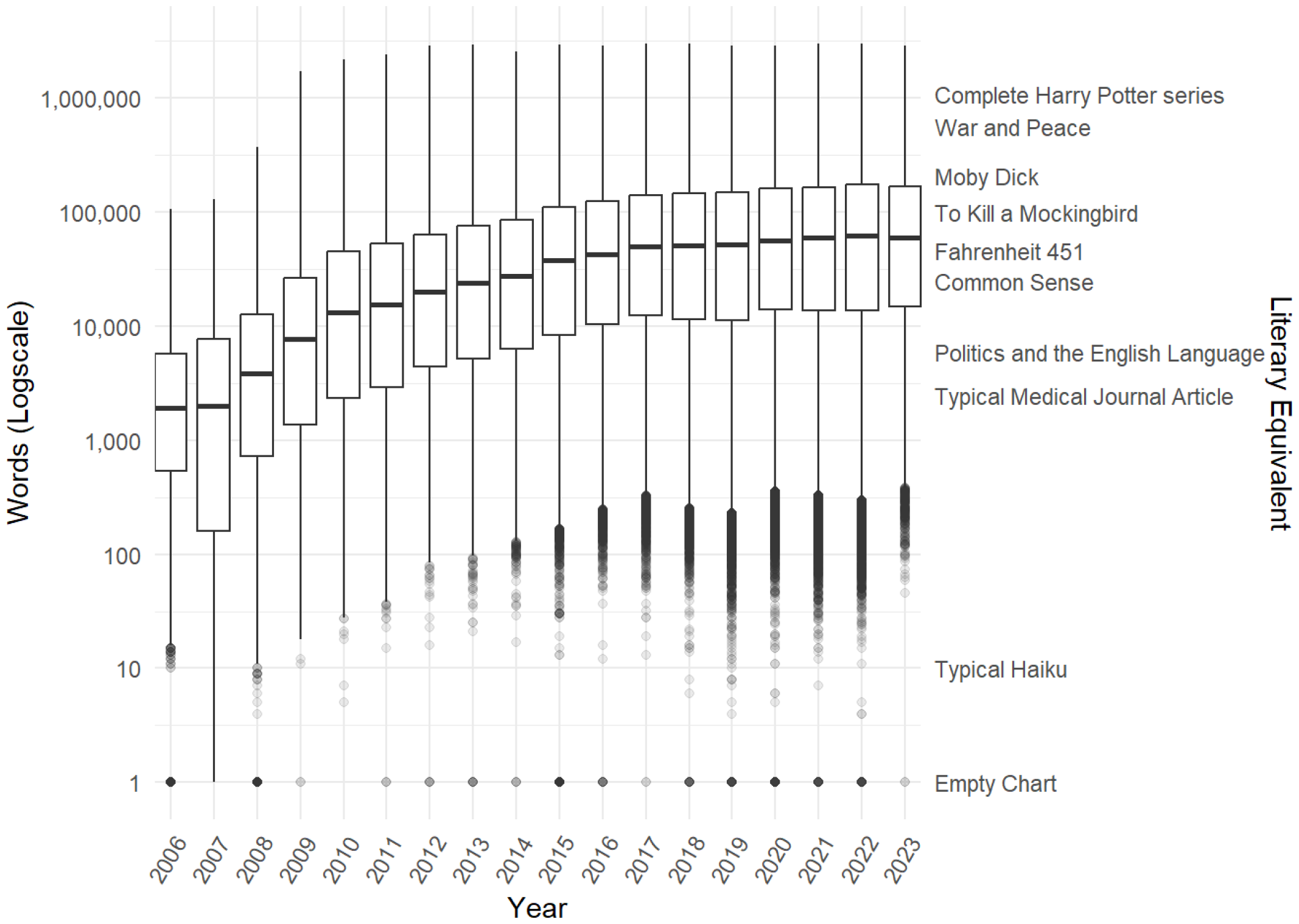
Total word counts for all notes available at an ED encounter, by year

Admission data was only available on the encounter level in 2009 and after. Note, word, and token counts were larger for patients admitted to general care units, who by 2022 had a median of 611 notes (IQR: 219 - 1,348) for 110,437 words (IQR: 34,672 - 247,559), and patients admitted to the ICU, who in 2022 had a median of 560 notes (IQR: 100 - 1,652) and 95,998 words (12,364 - 316,354).

Table 2 A and B evaluate the percentiles for aggregate word counts for notes available in 2022-2023, the most recent two years of data collection, as well as those available in encounters prior to 2010. Descriptive statistics for patient demographics and clinical characteristics with the corresponding note length are shown in Table 3.

**Table 2A:**
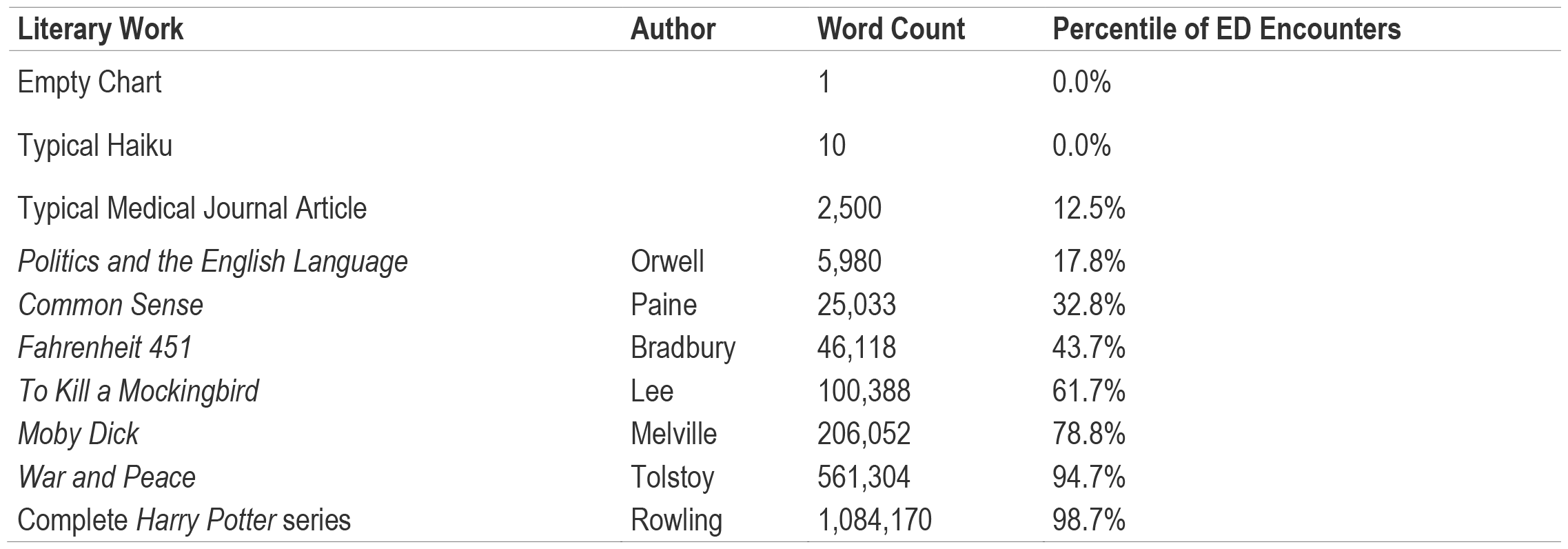
Available Note Text vs. Literary Works, 2022 and 2023.

**Table 2B:**
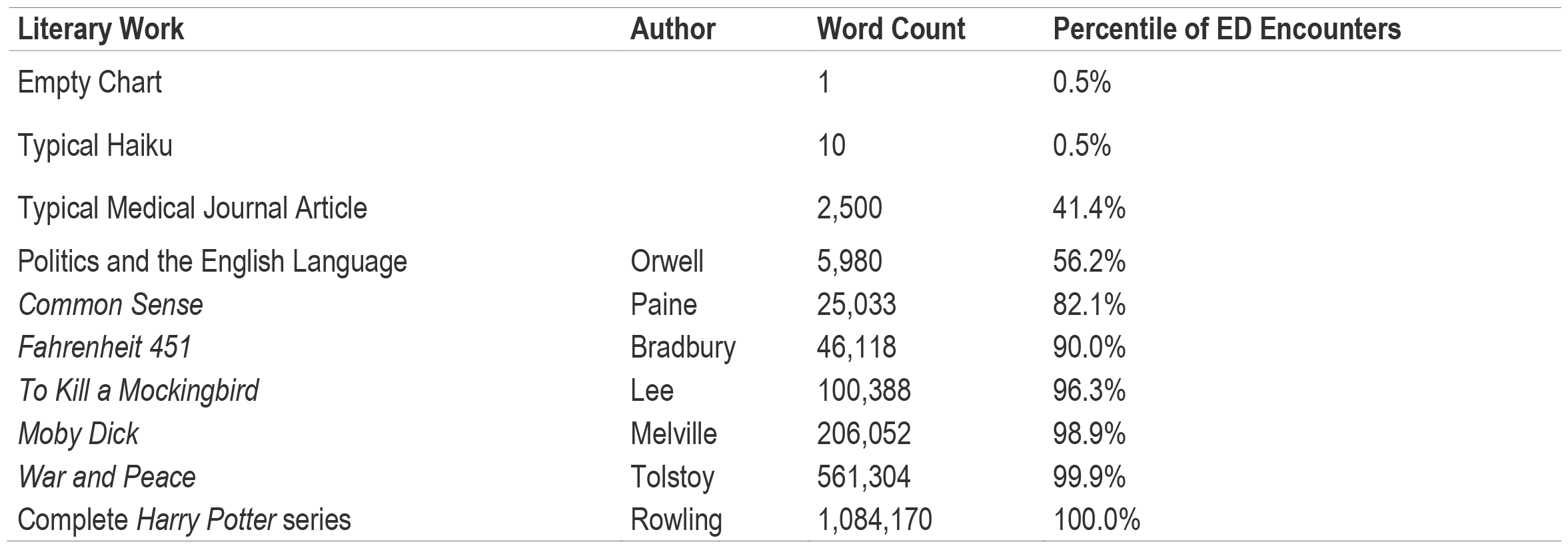
Available Note Text vs. Literary Works, 2010 and Prior.

| Literary Work | Author | Word Count | Percentile of ED Encounters |
| --- | --- | --- | --- |
| Empty Chart |  | 1 | 0.5% |
| Typical Haiku |  | 10 | 0.5% |
| Typical Medical Journal Article |  | 2,500 | 41.4% |
| Politics and the English Language | Orwell | 5,980 | 56.2% |
| *Common Sense* | Paine | 25,033 | 82.1% |
| *Fahrenheit 451* | Bradbury | 46,118 | 90.0% |
| *To Kill a Mockingbird* | Lee | 100,388 | 96.3% |
| *Moby Dick* | Melville | 206,052 | 98.9% |
| *War and Peace* | Tolstoy | 561,304 | 99.9% |
| Complete *Harry Potter* series | Rowling | 1,084,170 | 100.0% |

**Table 3:** Demographics Over Entire Study Period.

|  | n | Previous Note Count<br>(Median (IQR)) | Previous Note Words<br>(Median (IQR)) | Previous Note Tokens<br>(Median (IQR)) |
| --- | --- | --- | --- | --- |
| Overall | 730,968 | 200 (36 - 645) | 30,587 (5,145 - 106,757) | 51,368 (8,546 - 183,170) |
| **Age Group** |  |  |  |  |
| 18-34 | 222,811 | 61 (12 - 252) | 8,857 (1,507 - 38,566) | 14,551 (2,440 - 63,846) |
| 35-64 | 334,610 | 222 (52 - 648) | 33,911 (7,516 - 105,838) | 57,117 (12,565 - 181,362) |
| 65+ | 173,548 | 525 (165 - 1,143) | 87,959 (26,323 - 202,358) | 152,225 (45,272 - 354,277) |
| **Patient Biologic Sex** |  |  |  |  |
| Female | 394,084 | 250 (50 - 734) | 37,342 (7,201 - 118,482) | 62,782 (12,012 - 202,830) |
| Male | 336,869 | 148 (26 - 539) | 23,620 (3,513 - 92,225) | 39,537 (5,800 - 158,495) |
| Non-Binary Or Other | 16 | 21 (8 - 144) | 3,086 (1,484 - 22,274) | 4,602 (2,182 - 35,886) |
| **Patient Race** |  |  |  |  |
| American Indian Or Alaska Native | 8,238 | 127 (23 - 433) | 19,270 (3,085 - 72,274) | 32,189 (5,198 - 122,615) |
| Asian | 21,524 | 47 (9 - 249) | 6,814 (1,137 - 39,977) | 11,307 (1,820 - 67,108) |
| Black Or African American | 83,007 | 197 (40 - 642) | 29,894 (5,780 - 105,177) | 49,902 (9,567 - 178,764) |
| Native Hawaiian Or Other Pacific Islander | 1,356 | 108 (20 - 358) | 18,406 (2,577 - 58,724) | 30,611 (4,138 - 98,017) |
| White | 607,670 | 213 (39 - 670) | 32,716 (5,679 - 111,196) | 55,069 (9,438 - 191,026) |
| Not Recorded | 9,174 | 46 (10 - 228) | 6,870 (1,105 - 37,476) | 11,284 (1,775 - 62,989) |
| **Patient Ethnicity** |  |  |  |  |
| Hispanic/Latino | 34,823 | 92 (17 - 333) | 14,307 (2,274 - 55,591) | 24,001 (3,728 - 93,644) |
| Not Hispanic or Latino | 691,697 | 209 (38 - 666) | 32,008 (5,512 - 110,306) | 53,807 (9,152 - 189,265) |
| Not Recorded | 4,449 | 27 (8 - 195) | 3,298 (657 - 29,817) | 5,363 (1,064 - 50,349) |
| **Interpreter Needed for ED Encounter?** |  |  |  |  |
| Non-English Interpreter Needed | 18,330 | 79 (15 - 320) | 12,790 (1,891 - 57,924) | 21,488 (3,098 - 99,045) |
| No Interpreter Needed | 712,639 | 204 (37 - 654) | 31,199 (5,306 - 108,062) | 52,396 (8,818 - 185,288) |
| **Means of Arrival** |  |  |  |  |
| Family/Friend/Self | 532,880 | 176 (33 - 554) | 26,722 (4,762 - 89,782) | 44,721 (7,869 - 153,082) |
| EMS | 175,767 | 331 (57 - 1,010) | 52,536 (8,215 - 175,150) | 89,551 (13,824 - 304,734) |
| Helicopter | 5,094 | 32 (13 - 102) | 3,101 (1,377 - 13,073) | 5,366 (2,367 - 22,374) |
| Other or Not Recorded | 17,228 | 156 (18 - 646) | 25,846 (2,944 - 112,735) | 43,334 (4,818 - 193,412) |
| **Emergency Severity Index** |  |  |  |  |
| Level 1: Red | 5,482 | 102 (25 - 558) | 13,834 (2,478 - 94,216) | 23,496 (4,272 - 160,278) |
| Level 2: Pink | 154,528 | 279 (53 - 832) | 44,878 (7,773 - 146,720) | 76,372 (12,990 - 255,190) |
| Level 3: Yellow | 446,633 | 219 (44 - 671) | 33,589 (6,351 - 110,558) | 56,561 (10,570 - 189,534) |
| Level 4: Green | 113,328 | 87 (13 - 352) | 13,077 (1,905 - 54,090) | 21,648 (3,082 - 90,378) |
| Level 5: Blue | 9,724 | 69 (11 - 316) | 10,665 (1,577 - 49,053) | 17,493 (2,544 - 81,609) |
| Not Recorded | 1,274 | 84 (7 - 456) | 13,420 (1,614 - 73,897) | 22,170 (2,763 - 120,457) |
| **ED Disposition** |  |  |  |  |
| Discharge | 547,477 | 161 (29 - 518) | 24,175 (4,125 - 82,422) | 40,381 (6,784 - 140,003) |
| Admit | 172,031 | 400 (73 - 1,122) | 67,252 (11,384 - 202,597) | 116,004 (19,387 - 357,303) |
| Transfer | 11,461 | 401 (99 - 1,043) | 66,700 (15,072 - 178,329) | 113,187 (25,061 - 310,683) |
| **Has In-system Primary Care Provider?** |  |  |  |  |
| In-System PCP | 412,831 | 374 (121 - 924) | 58,850 (18,183 - 156,499) | 99,134 (30,292 - 269,338) |
| PCP Not Affiliated with ED System or No PCP | 318,138 | 54 (11 - 271) | 7,786 (1,278 - 42,948) | 12,997 (2,064 - 73,266) |

## Discussion

Numerous prior studies have examined the general state of information overload in the EHR and its contribution to burnout.^7–9,15,16^ Other work has specifically examined the phenomenon of note bloat, in which over time EHR notes have become longer and more repetitive,^12,17^ leading to stress among providers responsible for reading and creating notes.^18^ Less attention, however, has been paid to growth of the overall chart, and the related burden at the bedside for providers who routinely care for patients with whom they do not have an established prior relationship. Our findings demonstrate that the number of notes available to physicians in the EHR has significantly increased over the past decade, adding quantifiable evidence to the discussion on EHR-related physician burden. This highlights the increasing difficulty of an unassisted human “chart biopsy” task, especially in the ED. Emergency physicians often are responsible for caring for three or more new patients per hour,^17^ giving them an average time of 20 minutes for all patient care tasks: chart biopsy, face-to-face interview of the patient, physical examination, ordering and interpretation of test results, administration of therapies and procedures, discussions with consultants, writing discharge instructions, and counseling patients. Only a small fraction of these 20 minutes can be safely devoted to chart biopsy without affecting other critical patient care activities. While the time available for chart review has not increased, our results suggest that the chart biopsy task has drastically grown in magnitude since the inception of the EHR.

In this study, we used well known literary works to benchmark the volume of text data available in patient charts at the time of ED presentation. Prior to 2010, the task for a median patient was analogous to skimming a brief essay such as Orwell’s *Politics of the English Language* (6,000-words) to identify any potential salient points. This is difficult, but possible within a few minute window.

Today, a chart biopsy for the median patient is more analogous to skimming Fahrenheit 451 (a 46,000-word novel), while one in five patients arrive with a chart the size of *Moby Dick (*209,117 words). Skimming *Moby Dick* and identifying all possible health concerns for Captain Ahab is not a task a human can perform within the constraints of an ED visit. Inpatient and intensive care unit providers may have more time to conduct a chart review, but skimming *Moby Dick* is likely too time consuming and overwhelming in their setting as well. Furthermore, their task has grown to an even larger proportion: the average admitted patient has a chart over twice as long by word count as those examined during general ED visits.

These findings do not suggest that it is inherently wrong to store large amounts of text data within the EHR. The centralized, accessible storage of medical notes is a key benefit of EHR software, and studies have shown that even in the time-constrained ED setting, access to prior records improves physicians’ diagnostic ability.^19^ Since the time of early EHR adoption, there has been an understanding that there is a tension between the EHR’s function for documentation storage and its need to provide retrievable information to support clinical tasks.^20^ However, as the volume of textual data continues to grow, this tension has become irreconcilable without additional tools to filter and summarize information. In the current state, providers cannot read all or even a small fraction of the notes available to them during a chart biopsy and must use filters and other tools to sort information. While filters can help simplify views, these approaches risk missing critical pieces of data. The vast increase in magnitude of text stored points to the need for more sophisticated solutions to ensure patient safety. Prior studies have suggested strategies aimed at improving clinicians’ ability to curate or retrieve EHR data at the bedside,^21^ but these have not achieved widespread implementation. The potential application of LLMs offers a promising avenue for addressing the information overload challenge in EHRs.^22^ These models can be trained to generate concise and relevant summaries of patient notes, allowing physicians to quickly grasp essential information without sifting through extensive text data. Newer approaches in retrieval augmented generation with LLMs can retrieve relevant textual data, reducing factual errors in knowledge intensive tasks with the potential to reduce the cognitive load on the physician.^23^

One limitation of current large language models, however, is the constraint on the number of tokens that can be processed at once. For example, GPT-4 Turbo has a token input limit of up to 128,000 tokens. Most open-source LLMs have smaller token limits. These token limits pose challenges for summarizing large numbers of notes and highlight the need for research into strategies to both filter the input into such models and improve their ability to ingest large amounts of data for summarization, if they are to provide meaningful impact in improving care.

## Conclusion

This study quantifies the escalating challenge created by the volume of text data confronting providers during the course of caring for multiple simultaneous patients in the time-pressured environments of the ED and at admission to inpatient units. While the central storage of medical notes remains a critically important function for the EHR, our findings emphasize the urgency of addressing the resultant information overload for healthcare providers. While LLMs offer a potential venue for summarization, the volume of the summarization task exceeds current technological capacity. Further research and development in the field of natural language processing with LLMs will be crucial to assist healthcare providers in navigating the information-rich landscape of EHRs efficiently and effectively to provide optimal patient care.

## Data Availability

All data produced in the present study are available upon reasonable request to the authors, within institutional and legal constraints surrounding sharing of PHI.

## Acknowledgements

The authors would like to thank Joel Gordon, MD and Cherodeep Goswami for their support of this work and suggestions on the manuscript. None of the authors has any affiliation or financial involvement that conflicts with the material presented in this report.

**Appendix 1.** Notes, Words, and Tokens Over Time - All Years.

| Year | ED Encounters | Median (IQR) Previous Notes | Median (IQR) Previous Words | Median (IQR) Previous Tokens |
| --- | --- | --- | --- | --- |
| **All ED Encounters** |  |  |  |  |
| 2006 | 19,507 | 7 (3 - 18) | 1,894 (538 - 5,684) | 3,236 (960 - 9,724) |
| 2007 | 27,621 | 12 (3 - 38) | 2,004 (158 - 7,664) | 3,518 (328 - 13,288) |
| 2008 | 23,106 | 28 (9 - 79) | 3,830 (728 - 12,613) | 6,620 (1,251 - 21,818) |
| 2009 | 23,494 | 58 (15 - 171) | 7,620 (1,368 - 26,468) | 12,984 (2,278 - 45,684) |
| 2010 | 29,220 | 106 (25 - 308) | 13,179 (2,328 - 45,258) | 22,470 (3,907 - 78,514) |
| 2011 | 30,076 | 132 (31 - 385) | 15,552 (2,918 - 53,304) | 26,521 (4,890 - 92,960) |
| 2012 | 31,028 | 161 (39 - 452) | 19,978 (4,434 - 62,819) | 33,976 (7,272 - 109,097) |
| 2013 | 32,152 | 181 (42 - 515) | 24,089 (5,160 - 74,820) | 40,660 (8,516 - 128,984) |
| 2014 | 33,066 | 190 (45 - 543) | 27,322 (6,340 - 84,940) | 45,742 (10,432 - 146,021) |
| 2015 | 45,142 | 245 (58 - 677) | 37,323 (8,257 - 110,721) | 62,741 (13,616 - 190,807) |
| 2016 | 52,890 | 267 (64 - 732) | 42,753 (10,267 - 122,977) | 71,870 (16,886 - 211,348) |
| 2017 | 56,657 | 297 (73 - 791) | 49,885 (12,394 - 139,927) | 83,605 (20,314 - 240,979) |
| 2018 | 59,719 | 311 (75 - 841) | 51,147 (11,448 - 146,218) | 86,071 (18,834 - 251,322) |
| 2019 | 64,093 | 315 (75 - 858) | 51,500 (11,209 - 149,365) | 86,353 (18,411 - 256,802) |
| 2020 | 57,674 | 349 (90 - 926) | 56,254 (13,901 - 159,474) | 94,346 (22,923 - 274,413) |
| 2021 | 68,119 | 363 (92 - 950) | 59,393 (13,708 - 164,778) | 99,266 (22,418 - 282,588) |
| 2022 | 71,344 | 380 (93 - 1,008) | 61,591 (13,621 - 174,152) | 103,350 (22,375 - 298,983) |
| 2023 | 6,060 | 372 (101 - 963) | 59,677 (14,714 - 167,159) | 100,526 (24,300 - 287,460) |
| **ED Encounters Resulting in General Care Admission** |  |  |  |  |
| 2009 | 1,594 | 220 (64 - 543) | 35,126 (8,759 - 94,636) | 61,486 (15,165 - 166,759) |
| 2010 | 5,568 | 268 (82 - 626) | 40,920 (10,291 - 106,590) | 71,246 (17,752 - 190,243) |
| 2011 | 4,916 | 333 (100 - 800) | 49,579 (11,720 - 130,535) | 87,546 (20,490 - 236,774) |
| 2012 | 5,261 | 382 (115 - 921) | 53,138 (14,075 - 147,048) | 92,526 (24,277 - 263,638) |
| 2013 | 5,760 | 388 (111 - 946) | 58,192 (14,529 - 155,002) | 101,603 (24,633 - 274,833) |
| 2014 | 4,643 | 412 (116 - 1,025) | 67,970 (17,260 - 176,345) | 117,754 (29,468 - 312,853) |
| 2015 | 12,637 | 453 (130 - 1,100) | 74,948 (19,533 - 189,514) | 130,247 (32,994 - 334,208) |
| 2016 | 13,370 | 498 (150 - 1,214) | 85,956 (24,521 - 218,444) | 148,714 (41,665 - 385,562) |
| 2017 | 13,153 | 548 (166 - 1,288) | 100,460 (28,168 - 242,050) | 173,319 (47,427 - 426,692) |
| 2018 | 13,103 | 564 (162 - 1,356) | 100,763 (27,182 - 248,684) | 172,471 (45,854 - 435,658) |
| 2019 | 12,341 | 592 (172 - 1,384) | 104,148 (28,556 - 256,938) | 179,355 (47,829 - 451,177) |
| 2020 | 12,101 | 614 (186 - 1,474) | 109,270 (31,459 - 271,589) | 188,880 (53,057 - 474,857) |
| 2021 | 14,227 | 663 (201 - 1,596) | 117,626 (31,954 - 295,356) | 202,516 (53,780 - 514,496) |
| 2022 | 14,383 | 675 (202 - 1,636) | 117,906 (31,010 - 301,868) | 203,741 (52,154 - 529,416) |
| 2023 | 1,166 | 611 (219 - 1,348) | 110,437 (34,672 - 247,559) | 192,522 (59,471 - 430,562) |
| **ED Encounters Resulting in IMC/ICU Admission** |  |  |  |  |
| 2009 | 239 | 164 (30 - 534) | 24,780 (3,954 - 96,202) | 46,312 (6,842 - 167,132) |
| 2010 | 726 | 243 (60 - 659) | 36,260 (5,833 - 118,834) | 63,677 (10,076 - 212,672) |
| 2011 | 826 | 290 (54 - 930) | 42,423 (4,714 - 163,631) | 74,834 (8,138 - 293,147) |
| 2012 | 938 | 356 (60 - 1,012) | 51,970 (5,156 - 170,207) | 90,042 (9,366 - 310,728) |
| 2013 | 1,174 | 323 (56 - 952) | 46,968 (5,383 - 174,430) | 81,134 (8,824 - 313,865) |
| 2014 | 754 | 372 (72 - 988) | 61,493 (7,592 - 183,691) | 106,934 (12,580 - 328,589) |
| 2015 | 1,839 | 296 (46 - 952) | 50,200 (4,377 - 169,680) | 88,196 (7,486 - 302,946) |
| 2016 | 1,931 | 382 (66 - 1,150) | 66,827 (7,001 - 213,038) | 116,760 (11,836 - 380,311) |
| 2017 | 1,738 | 441 (74 - 1,279) | 80,226 (9,114 - 244,572) | 139,288 (15,590 - 436,836) |
| 2018 | 1,879 | 473 (78 - 1,370) | 84,802 (10,164 - 262,356) | 145,940 (17,294 - 470,318) |
| 2019 | 2,048 | 478 (95 - 1,344) | 81,170 (14,000 - 253,935) | 141,124 (24,123 - 443,804) |
| 2020 | 2,168 | 464 (96 - 1,262) | 82,230 (12,990 - 234,939) | 140,988 (22,153 - 416,106) |
| 2021 | 2,262 | 414 (68 - 1,306) | 70,307 (7,879 - 242,598) | 121,940 (13,888 - 425,681) |
| 2022 | 2,489 | 560 (100 - 1,652) | 95,998 (12,364 - 316,354) | 164,848 (20,869 - 554,417) |
| 2023 | 224 | 485 (98 - 1,102) | 98,991 (11,196 - 230,243) | 168,352 (18,434 - 414,983) |

*Note:*
IMC: Intermediate Care Unit; ICU: Intensive Care Unit

## References

1. Menachemi, N. & Collum, T. H. Benefits and drawbacks of electronic health record systems. Risk Manag. Healthc. Policy 4, 47–55 (2011).

2. Adler-Milstein, J. & Jha, A. K. HITECH Act Drove Large Gains In Hospital Electronic Health Record Adoption. Health Aff. 36, 1416–1422 (2017).

3. Bates, D. W., Saria, S., Ohno-Machado, L., Shah, A. & Escobar, G. Big data in health care: using analytics to identify and manage high-risk and high-cost patients. Health Aff. 33, 1123–1131 (2014).

4. Häyrinen, K., Saranto, K. & Nykänen, P. Definition, structure, content, use and impacts of electronic health records: a review of the research literature. Int. J. Med. Inform. 77, 291–304 (2008).

5. Stewart, W. F., Shah, N. R., Selna, M. J., Paulus, R. A. & Walker, J. M. Bridging the inferential gap: the electronic health record and clinical evidence. Health Aff. 26, w181–91 (2007).

6. Hilligoss, B. Dissecting the Pre-Handoff chart biopsy: Information seeking in the electronic health record. Proc. Am. Soc. Inf. Sci. Technol. 47, 1–10 (2010).

7. Babbott, S. et al. Electronic medical records and physician stress in primary care: results from the MEMO Study. J. Am. Med. Inform. Assoc. 21, e100–6 (2014).

8. Friedberg, M. W. et al. Factors Affecting Physician Professional Satisfaction and Their Implications for Patient Care, Health Systems, and Health Policy. Rand Health Q 3, 1 (2014).

9. Hill, R. G., Jr, Sears, L. M. & Melanson, S. W. 4000 clicks: a productivity analysis of electronic medical records in a community hospital ED. Am. J. Emerg. Med. 31, 1591–1594 (2013).

10. Wachter, R. M. Why Health Care Tech Is Still So Bad. The New York Times (2015).

11. Rosenbaum, L. Transitional Chaos or Enduring Harm? The EHR and the Disruption of Medicine. N. Engl. J. Med. 373, 1585–1588 (2015).

12. Rule, A., Bedrick, S., Chiang, M. F. & Hribar, M. R. Length and Redundancy of Outpatient Progress Notes Across a Decade at an Academic Medical Center. JAMA Netw Open 4, e2115334 (2021).

13. Steinkamp, J., Kantrowitz, J. J. & Airan-Javia, S. Prevalence and Sources of Duplicate Information in the Electronic Medical Record. JAMA Netw Open 5, e2233348 (2022).

14. tiktoken. How to count tokens with tiktoken https://github.com/openai/openai-cookbook/blob/main/examples/How_to_count_tokens_with_tiktoken.ipynb (2023).

15. Kroth, P. J. et al. The electronic elephant in the room: Physicians and the electronic health record. JAMIA Open 1, 49–56 (2018).

16. Kroth, P. J. et al. Association of Electronic Health Record Design and Use Factors With Clinician Stress and Burnout. JAMA Netw Open 2, e199609 (2019).

17. Reznek, M. A., Michael, S. S., Harbertson, C. A., Scheulen, J. J. & Augustine, J. J. Clinical operations of academic versus non-academic emergency departments: a descriptive comparison of two large emergency department operations surveys. BMC Emerg. Med. 19, 72 (2019).

18. Apathy, N. C., Rotenstein, L., Bates, D. W. & Holmgren, A. J. Documentation dynamics: Note composition, burden, and physician efficiency. Health Serv. Res. 58, 674–685 (2023).

19. Ben-Assuli, O., Sagi, D., Leshno, M., Ironi, A. & Ziv, A. Improving diagnostic accuracy using EHR in emergency departments: A simulation-based study. J. Biomed. Inform. 55, 31–40 (2015).

20. Embi, P. J. et al. Computerized provider documentation: findings and implications of a multisite study of clinicians and administrators. J. Am. Med. Inform. Assoc. 20, 718–726 (2013).

21. Semanik, M. G. et al. Impact of a problem-oriented view on clinical data retrieval. J. Am. Med. Inform. Assoc. 28, 899–906 (2021).

22. Clusmann, J. et al. The future landscape of large language models in medicine. Commun. Med. 3, 141 (2023).

23. Ram, O. et al. In-Context retrieval-augmented language models. Trans. Assoc. Comput. Linguist. 11, 1316–1331 (2023).

